# EXPERIENCE OF THE USE OF VACUUM EXTRACTOR IN DELIVERY ASSISTANCE

**DOI:** 10.1101/2025.09.26.25336777

**Authors:** Armando Alberto Moreno Santillán, Leidy Marcela Martínez Adame, Brenda Cunningham

## Abstract

**Background:** The vacuum extractor is a tool that, through controlled vacuum generation, produces negative pressure on the fetal head, allowing traction, flexion, rotation, and extraction. It is currently considered a safe and effective alternative for assisted vaginal delivery.

**Objective:** To describe the indications, technical application parameters, complications, and success of the use of the vacuum extractor in childbirth assistance.

**Materials and Methods:** A retrospective, observational, and descriptive study was conducted in patients with singleton term pregnancies, in whom delivery was assisted with a rigid-cup vacuum extractor over a five-year period. Indication, technical parameters, complications, and success of the procedure were evaluated and recorded.

**Results:** The rigid-cup vacuum extractor was used in 105 patients. The main indications for its application were maternal exhaustion (n=50; 47.6%), prolonged second stage of labor (n=26; 24.8%), and risk of fetal distress (n=10; 9.5%). Successful extraction of the fetal head (defined as successful vacuum delivery) was achieved in 103 cases (98.1%). The most frequent maternal complication was first-degree vaginal tear (n=13; 12.4%), while the most common neonatal finding was caput succedaneum (n=20; 19.0%).

**Conclusions:** The use of a rigid-cup vacuum extractor is a useful tool, with a high success rate and a low risk of maternal or neonatal injury.

## BACKGROUND

The first description of the vacuum in history was made by James Younge in London in 1706, who used metal and glass cranial cups to shorten complicated labor. Later, in 1849, James Simpson in Edinburgh published the article *“The suction tractor or a new mechanical power as a substitute for forceps in difficult labors,”* in which he described the use of a device called the “aero-extractor,” consisting of a rubber cup connected to a suction pump instrument that generated a partial vacuum.^1^ In 1890, MacCahey, in the United States of America, designed a new suction cup he called the “atmospheric extractor,” which consisted of a rubber cup with a rigid handle that facilitated fetal traction.^1-^□ In 1948, Cunnigham, in Australia, developed a rubber cup connected to a series of bottles with a mercury manometer and an electric aspirator; however, this device was only tested in stillborn fetuses. Four years later, Malstrom in Sweden introduced his “vacuum extractor” as a new obstetric instrument and reported his experience in a cohort of 192 patients.^1, 2^,□ From Malstrom’s vacuum model, modifications and improvements were developed, leading to the modern vacuum extractor used today. ^□, □^

The vacuum extractor is a device that facilitates traction, flexion, and, in certain cases, rotation of the fetal head, and is therefore recognized as an alternative to forceps and spatulas. Currently, it is even considered the instrument of choice for operative vaginal delivery.^2^□□ In the United States, approximately 5% of births are operative, and the vacuum extractor is used almost four times more frequently than forceps.□ In other countries, such as Chile, the vacuum extractor has accounted for 28.6% of all operative vaginal deliveries. □

The use of the vacuum extractor is associated with a lower rate of cesarean sections, fewer immediate maternal complications, and faster postpartum recovery, making it a more advantageous alternative compared with forceps from the maternal perspective. It is also linked to a .^2-^□ ^−1^□ The frequency of operative vaginal deliveries varies between countries and hospitals, largely depending on the criteria of each obstetric school.□ In recent years, a decline in the use of forceps has been observed, while the use of vacuum has shown a relative increase, possibly due to its being considered less traumatic.^11–13^

Several international studies support the safety and efficacy of the vacuum extractor. A lower incidence of maternal and neonatal trauma compared with forceps has been reported, as well as success rates close to 95%, even in unfavorable presentations.^1^□ ^−1^□ In Mexico, although there are studies such as that by Moreno-Santillán et al., which compared forceps and vacuum,^1^□ and that by Arvizu et al., on perineal tears,^1^□ national evidence remains scarce. In this context, the objective of the present study is to evaluate the indications, technique, and complications of the Kiwi ProCup rigid-cup vacuum extractor, providing local data to strengthen the existing evidence.

## MATERIALS AND METHODS

A retrospective, observational, and descriptive study was conducted in patients with singleton term pregnancies who required operative vaginal delivery, attended at the obstetric surgery unit of the High Specialty Medical Unit, Hospital of Gynecology and Obstetrics No. 4 “Luis Castelazo Ayala,” in whom delivery was assisted using a vacuum extractor (rigid-cup Kiwi type). The study period spanned from April 1, 2017, to May 31, 2022.

Inclusion criteria were women in the second stage of labor in whom a rigid-cup vacuum extractor was applied, with singleton term pregnancy (37 to 41 weeks of gestation), meeting the following conditions for application: cephalic presentation, ruptured membranes, obstetric analgesia, and maternal or fetal indication for operative vaginal delivery.

For the descriptive univariate analysis, quantitative variables are presented as measures of central tendency and dispersion, according to their distribution, and qualitative variables as percentages. Statistical analysis was performed using RStudio software, version 4.1.0 © 2009–2021.

This study was reviewed and approved by the Local Research Committee of the High Specialty Medical Unit No. 4 “Luis Castelazo Ayala” and was carried out in accordance with the General Health Law on Research. It was classified as “minimal risk,” as it was conducted solely through the review of documentary records.

## RESULTS

The rigid-cup vacuum extractor was applied to 105 women, whose main characteristics are presented in Table 1.

**Table 1.** General characteristics of the study population:

| Variable | Description |
| --- | --- |
| Maternal age (mean ± standard deviation) | 26.5 ± 5.1 years |
| Gestational age (mean ± standard deviation) | 39.1 ± 0.9 weeks |
| Primigravidas (number and percentage) | 60 (57.2%) |
| Secundigravidas (number and percentage) | 32 (30.5%) |
| Multigravidas (number and percentage) | 13 (12.4%) |
| Maternal conditions |  |
| · Gestational diabetes (number and percentage) | 7 (6.7%) |
| · Previous cesarean section (number and percentage) | 2 (1.9%) |
| · Hypothyroidism (number and percentage) | 2 (1.9%) |
| · Gestational hypertension (number and percentage) | 1 (0.9%) |
| · Grade 3 obesity (number and percentage) | 1 (0.9%) |

The indications for application were maternal exhaustion, prolonged second stage, risk of fetal distress, need to shorten the second stage due to previous cesarean section, fetal bradycardia, placental abruption, fetal tachycardia, tachysystole, and maternal heart disease (Table 2).

**Table 2.** Indications for vacuum extractor application:

| Indication | Number of patients and percentage |
| --- | --- |
| Maternal exhaustion | 50 (47.6%) |
| Prolonged second stage | 26 (24.8%) |
| Risk of fetal distress | 10 (9.5%) |
| Need to shorten the second stage | 7 (6.7%) |
| Fetal bradycardia | 5 (4.8%) |
| Placental abruption (normally inserted) | 3 (2.9%) |
| Fetal tachycardia | 2 (1.9%) |
| Tachysystole | 1 (0.9%) |
| Maternal heart disease | 1 (0.9%) |

Regarding the type of analgesia used for the placement of the vacuum extractor, 96 patients (91.4%) received epidural block and 9 (8.6%) received pudendal nerve block. In all cases (100%), the procedure was performed with complete cervical dilation, ruptured membranes, an empty bladder achieved by catheterization, and application of the cup at the flexion point of the fetal head. A mediolateral episiotomy was performed in 96 patients (91.4%), while in 9 cases (8.6%) no additional techniques to widen the birth canal were required.

Regarding the level of vacuum placement, in 89 cases (84.8%) it was performed at the third Hodge plane, and in 16 cases (15.2%) at the second plane. With respect to fetal position, most applications were performed in the occiput anterior position (n=74; 70.5%), followed by left occiput anterior (n=11; 10.5%), right occiput anterior (n=7; 6.7%), occiput posterior (n=10; 9.5%), right occiput transverse (n=2; 1.9%), and left occiput transverse (n=1; 0.9%).

Successful extraction of the fetal head with vacuum (defined as successful vacuum delivery) was achieved in 103 cases (98.1%). To accomplish this, a mean vacuum pressure of 542.9 ± 22.2 mmHg was required, with an average traction force of 8.3 ± 0.6 kg. In the remaining two cases (1.9%), in which birth could not be achieved with vacuum, Salinas-type forceps were required to complete fetal extraction. The main neonatal characteristics of the 105 newborns are summarized in Table 3.

**Table 3.** General characteristics of the newborns:

| Variable | Description |
| --- | --- |
| Female sex (number and percentage) | 45 (42.9%) |
| Weight (mean ± standard deviation) | $3023.7 \pm 322.7$ g |
| Apgar score at 1 minute (mean ± standard deviation) | $8.16 \pm 0.7$ |
| Apgar score at 5 minutes (mean ± standard deviation) | $8.9 \pm 0.3$ |
| Length (mean ± standard deviation) | $49.6 \pm 1.5$ cm |
| Head circumference (mean ± standard deviation) | $33.5 \pm 1.3$ cm |
| Delayed umbilical cord clamping (number and percentage) | 93 (88.6%) |
| Immediate skin-to-skin contact (number and percentage) | 95 (90.5%) |

Regarding the area to which the newborns were admitted, 82 (78%) were assigned to rooming-in, 22 (20.1%) to the intermediate care nursery, and one to the neonatal intensive care unit due to perinatal asphyxia. Of the 22 newborns admitted to the intermediate care nursery, 16 were due to transient tachypnea of the newborn, 2 for suspected neonatal infection, 2 for low birth weight, 1 for high birth weight, and 1 for tachypnea.

Maternal complications associated with the application of the vacuum extractor included first-degree vaginal laceration in 13 patients (12.4%), second-degree perineal laceration in one case (0.9%), third-degree laceration in one case (0.9%), and fourth-degree laceration in one case (0.9%).

As for neonatal complications, caput succedaneum was documented in 20 newborns (19.0%), while cephalohematoma and skin lacerations were observed in one case each (0.9%).

## DISCUSSION

Given that national evidence on the use of the vacuum extractor is limited and derived from only a few studies, this work evaluated its indications, technique, and complications using the Kiwi ProCup rigid-cup model. In 105 operative vaginal deliveries in singleton term pregnancies, a high success rate was observed, with the main indications being maternal exhaustion, prolonged second stage, and suspected fetal distress. Most procedures were performed under epidural analgesia, with cup placement at the third Hodge plane and in the occiput anterior position. Maternal complications were mild, mainly first-degree vaginal lacerations, while in newborns, caput succedaneum was predominant; Apgar scores were favorable, and more than three-quarters were assigned to rooming-in.

Several publications have documented the increasing use of vacuum or obstetric suction devices in different developed countries. For example, Curtin reported a rise in its use in the United States since 1992, with vacuum being employed twice as often as forceps starting in 1997.^1^□ ^−2^□ At Southmead Hospital in Bristol, United Kingdom, where 5,000 births are attended annually, the rate of vacuum-assisted deliveries is 8^−^10% per year.^21^ In Canada, the rate of vacuum-assisted deliveries increased from 0.6% in 1980 to 10.6% in 2001, while forceps deliveries decreased from 21.2% in 1980 to 6.8% in 2001. However, in other regions of the world, such as Latin America, the use of vacuum has not been fully implemented, with high cesarean section rates. This is the case in Chile, Brazil, Paraguay, and our country, where cesarean rates continue to increase, even exceeding 50% in the private sector and some public institutions—far above the 10–15% recommended by World Health Organization guidelines. In our hospital, during 2018, of a total of 12,596 births, only 50.19% (6,322) were vaginal deliveries, while 49.5% (6,236) were cesarean sections.^22^

One of the largest cohort studies on vacuum-assisted delivery was conducted at Dalhousie University, including 1,000 participants. In that study, a success rate of 87% was reported for vacuum application, with complementary use of forceps required in 9.8% of cases and a subsequent cesarean section rate of only 2%.^23^ In contrast, in Mexico there is a lack of recent statistics on the use of the vacuum extractor, which highlights the value of our institutional experience. In our series, a success rate of 97.7% was documented in 90 patients, with complementary use of Salinas-type forceps in only two cases (2.2%) and no need for conversion to cesarean section in any of them. These results are consistent with those described by Falcone (2025)^1^□ and Goetzinger et al. (2008)^2^□, who reported success rates between 90% and 98%, depending on operator expertise and proper case selection. This concordance reinforces the external validity of our findings and positions the vacuum extractor as a reliable and effective tool in the current obstetric context.

The main indications identified in this study—maternal exhaustion, prolonged second stage, and suspected fetal distress—are consistent with international clinical recommendations. The ACOG Practice Bulletin No. 219, 2020, establishes as valid indications for operative vaginal delivery the prolongation of the second stage of labor, fetal compromise, and the need to shorten the second stage due to maternal exhaustion.^2^□ Similarly, Tonismae (2023) and Hook and Damos (2008) include maternal exhaustion, fetal tachycardia or bradycardia, and prolonged second stage as appropriate clinical scenarios for the use of vacuum.^2^□ ^−2^□ In our cohort, maternal exhaustion was the most frequent indication (47.6%), followed by prolonged second stage (24.7%) and suspected fetal distress (9.5%), reflecting a clinical application consistent with internationally established guidelines and demonstrating medical practice aligned with the current literature.^2^□ ^−2^□

It is essential to consider not only the clinical indications for the use of the vacuum extractor but also the strict adherence to technical requirements and the correct application of the device, as these factors significantly reduce maternal and fetal complications and increase the success rate. In this regard, the study by Dr. Aldo Vacca, which included 119 patients, highlighted the importance of precise cup placement on the fetal head and the use of devices that allow monitoring of the traction force applied.^3^□ According to his research, limiting traction to a maximum of 11.5 kg helps prevent fetal injuries and also reduces severe maternal complications, such as anal sphincter damage, achieving vaginal deliveries in up to 80% of cases. In our series, we observed results consistent with these recommendations. Effective traction was achieved with an average force of 8.3 ± 0.6 kg and a mean vacuum pressure of 542.9 ± 22.2 mmHg. Extraction was successful on the first attempt in 46 cases (51.1%), on the second in 22 cases (24.4%), on the third in 15 cases (16.6%), on the fourth in 3 cases (3.3%), and on the fifth in 2 cases (2.2%). These results highlight the advantages of the Kiwi silicone rigid-cup vacuum extractor, which allows more precise control of technical parameters and ensures greater safety during operative vaginal delivery.

One of the drawbacks previously associated with the use of vacuum was its lower success rate compared with forceps; however, this concern has been overcome thanks to improvements in obstetric suction devices, particularly with the Kiwi rigid-cup vacuum. Unlike soft cups, the rigid cup shows higher success rates without increasing the risk of fetal scalp lacerations, with similar success outcomes to the silicone rigid cup, in addition to a lower rate of maternal injuries compared with forceps.^2^□ ^−2^□ In our study, the observed complications, such as caput succedaneum (19%) and first-degree vaginal lacerations (12.4%), were consistent with previous reports, such as the study conducted by Abbas (2021), which compared operative and spontaneous deliveries, showing that these injuries are the most common in vacuum-assisted births, generally being mild and self-limiting.^31^ Moreover, it was highlighted that severe complications such as cephalohematoma or subgaleal hemorrhage are rare when proper technical criteria are followed.

One of the main strengths of this study is that it provides current and relevant evidence on the use of the vacuum extractor in our clinical setting, in a context where health systems are seeking safe modalities to reduce unnecessary cesarean section rates. Our findings support the use of vacuum as an effective and low-risk alternative to complete delivery, with minimal maternal and fetal complications. This is consistent with the Chilean review that compared 12 randomized controlled trials, concluding that the use of vacuum is associated with a lower rate of severe maternal injuries compared with forceps, without increasing perinatal mortality or other significant fetal complications (Cuevas et al., 2007).□

Among the limitations, the retrospective design of the study stands out, which implies reliance on clinical records and a potential information bias. In addition, no comparative group with forceps was included, nor were long-term neonatal outcomes evaluated. Variables such as the individual operator’s experience, which could have influenced the success rate and frequency of complications, were also not analyzed. Nevertheless, these findings are supported by the adequate sample size, the homogeneity of the population, and the technical standardization in the application of the Kiwi ProCup device.

The results of this study reinforce the role of the rigid-cup vacuum extractor as a safe and effective tool in childbirth assistance, particularly in contexts where reducing cesarean section rates is a priority without compromising maternal and fetal safety. Proper selection of indications, adherence to technical requirements, and the use of devices that allow monitoring of parameters such as traction force and applied pressure are essential to achieve successful outcomes with a low complication rate. Although further prospective and comparative studies including forceps or other methods are required, our findings contribute to strengthening the clinical evidence in favor of vacuum as a valid and up-to-date alternative in obstetric practice. The Kiwi-type vacuum extractor, when correctly indicated and applied, is a safe and effective alternative for completing vaginal delivery in selected clinical scenarios.

## CONCLUSIONS

The use of the rigid-cup vacuum extractor has proven to be a useful tool, easy to apply, with a high success rate and a low risk of maternal or fetal injuries.

## Data Availability

All data produced in the present work are contained in the manuscript

